# Exhaled Mycobacterium tuberculosis predicts incident infection in household contacts

**DOI:** 10.1101/2021.11.06.21266008

**Authors:** CML Williams, A Muhammad, B Sambou, A Bojang, A Jobe, G Daffeh, O Owolabi, D Pan, M Pareek, MR Barer, JS Sutherland, P Haldar

**Author notes:** Joint Senior Author (MR Barer, JS Sutherland, P Haldar). **Corresponding Author:** Dr Caroline Williams. Department of Respiratory Sciences, University of Leicester, University Road, Leicester UK. **Author Contributions:** CMLW: study conceptualization, data collection, laboratory processing of samples, data interpretation, statistical analysis and drafting of manuscript. AM: statistical analysis, manuscript drafting and review. BS, AB & GD: laboratory processing of samples. AJ: data collection. OO: data collection, manuscript drafting and review. DP: statistical analysis, manuscript drafting and review. MP: data interpretation, manuscript drafting & review. MRB: study conceptualization, manuscript drafting and review. JSS & PH: project supervision, study conceptualization, data interpretation, statistical analysis, manuscript drafting and review.

## Abstract

**Background:** Halting transmission of *Mycobacterium tuberculosis* (Mtb) by identifying infectious individuals early is key to eradicating Tuberculosis (TB). Here we evaluate face mask sampling as a tool for stratifying infection risk in household contacts of pulmonary TB (pTB).

**Methods:** Forty-six sputum positive pTB patients in The Gambia (Aug 2016-Nov 2017) consented to mask sampling prior to commencing treatment. Incident Mtb infection was defined in their 181 household contacts as QuantiFERON (QFT) conversion or an increase in Interferon-ƴ release of ≥ 1IU/ml, 6 months after index diagnosis. Multilevel mixed-effects logistical regression analysis with cluster adjustment by household was used to identify predictors of incident infection.

**Findings:** Mtb was detected in 91% of pTB mask samples with high variation in IS6110 copies (5.3 ×10^2^ to 1.2 ×10^7^). A high mask Mtb level (≥20,000 IS6110 copies) was observed in 45% of cases and independently associated with increased likelihood of incident Mtb infection in contacts (AOR (95%CI) 3.20 (1.26 - 8.12), p=0.01), compared with cases having low/negative mask Mtb levels. Mask Mtb level was a better predictor of incident Mtb infection than sputum bacillary load, chest radiographic characteristics or sleeping proximity.

**Interpretation:** Mask sampling offers a highly sensitive and non-invasive tool to support both diagnosis of pTB and stratification of individuals who are most infectious. Our findings have the potential to revolutionise contact screening strategies and outbreak management in high TB burden settings and is of urgent public health importance.

## Introduction

Tuberculosis (TB) remains a major global health challenge^1^ which has been compounded by COVID-19 and its effect on health services worldwide^2^. The causative organism, *Mycobacterium tuberculosis* (Mtb), is reliant on airborne transmission for survival and so, identifying those individuals who are emitting infectious bacilli is key to global eradication^3^.

The World Health Organisation (WHO) advocates screening close contacts of pulmonary TB (pTB) cases to identify recently infected individuals as part of the TB elimination strategy^4^. For high burden settings in particular, this dovetails with active case finding programmes to identify subclinical TB, which is increasingly recognised to be a potentially important contributor to the overall burden of transmission.^5^

However, in these settings systematic screening is severely constrained by insufficient resources^6^. To overcome this, tools that can rapidly identify the most infectious individuals could enable more focussed contact screening pathways to be developed. However, traditional clinical markers of infectivity that include bacillary burden in sputum, extent of disease (particularly cavitation) on chest x-rays and the presence of cough have repeatedly been shown to be unreliable in the assessment of transmission at the individual level^7–11^.

Studies to date suggest direct sampling of infectious aerosolised bacilli from infected individuals using the Cough Aerosol Sampling System (CASS) can better discriminate between pTB patients with high and low levels of infectiousness than traditional markers^8^. CASS detects colony forming unit (CFU) bacilli captured in an Andersen sampler following two five-minute bouts of coughing. Although effective, it is a relatively demanding technique, requiring carefully calibrated apparatus, trained personnel and access to a biocontainment level 3 laboratory.

We have developed face mask sampling (FMS) as an alternative method for quantifying bacilli exhaled by pTB patients, in association with their natural breathing pattern and coughing. The approach is simple, non-invasive and applicable in any setting where a mask can be worn. We have demonstrated the capacity of FMS to reveal individual patterns of Mtb emission over 24 hours and shown that these are often dissociated from concomitant sputum bacillary output. As a screening tool for diagnosing pulmonary TB, our studies indicate that FMS can detect active TB more consistently and with greater sensitivity than conventional sputum analysis^12,13^.

In this prospective cohort study we extend our evaluation of FMS to investigate its utility for stratifying infectiousness of pTB. We have hypothesised that emitted bacillary genomic signals detected by FMS could be correlated with household contact transmission rates. The work was conducted in household contacts of pTB patients, the high TB burden setting of The Gambia, where both *M. africanum* and *Mtb* cause clinical disease^14^.

## Methods

### Study population and design

Sputum acid-fast bacilli (AFB) smear positive pulmonary TB patients were recruited between August 2016-November 2017, in the Greater Banjul Area of The Gambia in West Africa. Recruitment took place at either the MRC TB clinic (Fajara) or four local health centres (Brikama, Fajikunda, Jammeh Foundation for Peace Hospital and Serrekunda). Patients aged ≥18 years with at least 3 adult household members in close contact for at least 3 months were eligible for enrolment to the study. Recruited patients provided a 60 minute mask sample (see below) prior to treatment. A baseline chest X-ray (CXRs) was performed in all patients and scored for disease extent and cavitation (appendix).

Household contacts of the recruited pulmonary TB cases were assessed at baseline for signs of active TB with detailed medical history, clinical examination, CXR and, where possible, sputum analysis. Household contacts in whom active disease was excluded were eligible for the study if they had not been treated for TB in the previous 12 months, lived with the index TB patient and willing to take part. All recruited household contacts were tested for HIV at baseline and had blood taken for QuantiFERON TB Gold-in-Tube testing (QFT, Qiagen, Germany) at enrolment and 6 months. No treatment was given to patients identified with LTBI during the course of the study.

In addition to routine demographic and clinical data for both TB cases and contacts, specific information about the whole household was collected, including the number and age of all people living in each compound and sleeping proximity of each person to the TB case.

### Sputum processing

Processing of the initial screening sputum sample for acid-fast bacilli (AFB) analysis was performed at the community clinic. All other samples for the TB cases were processed at the microbiology laboratory at the MRC Unit in Fajara. These included sputum AFB smear microscopy, Xpert MTB/RIF assay (Cepheid, USA) and liquid culture (BACTEC MGIT 960, Becton Dickinson, USA). Mycobacterial species was determined by spoligotyping^15^ and radiologic extent of disease was graded on a four-category ordinal scale independently by two members of the research team (see appendix for details).

### Face mask sampling and processing

Index cases wore a modified face mask containing a gelatine sampling matrix for 1 hour under direct observation from which Mtb DNA was extracted and quantified, as previously described^12^. Briefly, exposed gelatine from the face mask was dissolved in sodium hydroxide (1·5 mL of 2% w/v), neutralised with 190 μL 4 mol/L hydrochloric acid, centrifuged at 13,400 × g for 10 min, then the pellet was re-suspended in TE buffer (comprising the pH buffer Tris and the cation chelator EDTA and stored at −80°C. Cells were subsequently disrupted by bead-beating and DNA extraction based on the methods of Reddy and colleagues^16^. Bacillary burden was assayed by IS6110-directed PCR^17^.

### QuantiFERON analysis

At baseline and 6 months, whole blood was collected from the recruited TB contacts and tested for immunoreactivity to Mtb antigens using the QuantiFERON TB Gold In Tube assay (1 ml per tube for TB-Ag, NIL and PHA) in accordance with manufacturer’s instructions. Data was analysed using the manufacturer’s recommended positive cut-off of ≥ 0.35 IU/ML and ≥ 25% of the NIL response after subtracting the NIL from each antigen-specific response

### Statistical Analysis

Our primary exposure variable was mask output of the index case within the household and correlation of this with infection in their household contact (HHC). To evaluate a relationship between the quantity of IS6110 captured by mask sampling and QFT conversion, we divided mask output into two groups after first looking at the trend in risk of QFT conversion by finely divided categories of genomic copies, as described by Jones-Lopez and colleagues using CASS.^8^ On this basis, we defined two groups: (1) ≥ 20,000 copies and (2) < 20,000 copies or negative.

Our primary outcome measure was incident Mtb infection within the household contact defined by QFT conversion from negative to positive. Our secondary outcome measure used a quantitative increase in the QFT reading from baseline to 6 months of ≥ 1 IU/ml, regardless of baseline IFN-ƴ result. This secondary measure was included to overcome the inherent risk of missed seroconversion events in contacts arising from delayed index diagnosis. A threshold increment of ≥ 1 IU/ml was chosen as studies have suggested that an IFN-ƴ level of < 1 IU/ml is less likely to represent a significant infection event^18,19^ and that significant changes in the quantitative value of QFT are associated with a higher incidence of TB disease progression^20,21^.

We calculated a Negative Predictive Value (NPV) of FMS to predict incident infection (QFT conversion and IFN-ƴ increase of ≥ 1 IU/ml) with an average transmission rate to household contacts of 29%.^22^

We calculated descriptive statistics of clinical and demographic characteristics of contacts of index cases and compared differences between subgroups with those exposed to high mask output and low/negative mask output. (see appendix for details) To identify factors associated with new Mtb infection in contacts, we performed multilevel mixed-effects logistic regression, adjusting for clustering within households. Parameters achieving a statistical threshold of p<0.1 in univariable analysis were retained for inclusion to the multivariable multilevel mixed effects logistic regression model (Table S4 in supplemental data). All statistical analyses were performed using STATA version 13.1 (StataCorp LP, USA). Results are presented as unadjusted and adjusted odds ratios (OR and AOR) with 95% confidence intervals (CI).

### Ethics Statement

Written, informed consent was obtained from all participants prior to sample collection. Ethical approved was provided by the Gambia Government/MRC joint ethics committee (ref: SCC 1486v2).

## Results

Between February 2017 and May 2018, we screened 64 sputum AFB smear positive pulmonary TB patients and enrolled 50 participants (Figure 1). Four index cases and their household contacts (HHC) were later withdrawn from study analysis because all HHC were lost to follow up or had missing results. In total 181 (85%) of 217 household contacts recruited from households of the 46 index cases provided serial QFT data for analysis (Figure 1). There was no difference in baseline characteristics between TB cases and household contacts that were included and excluded from the study (Table S2 & S3 in supplemental data).

**Figure 1.**
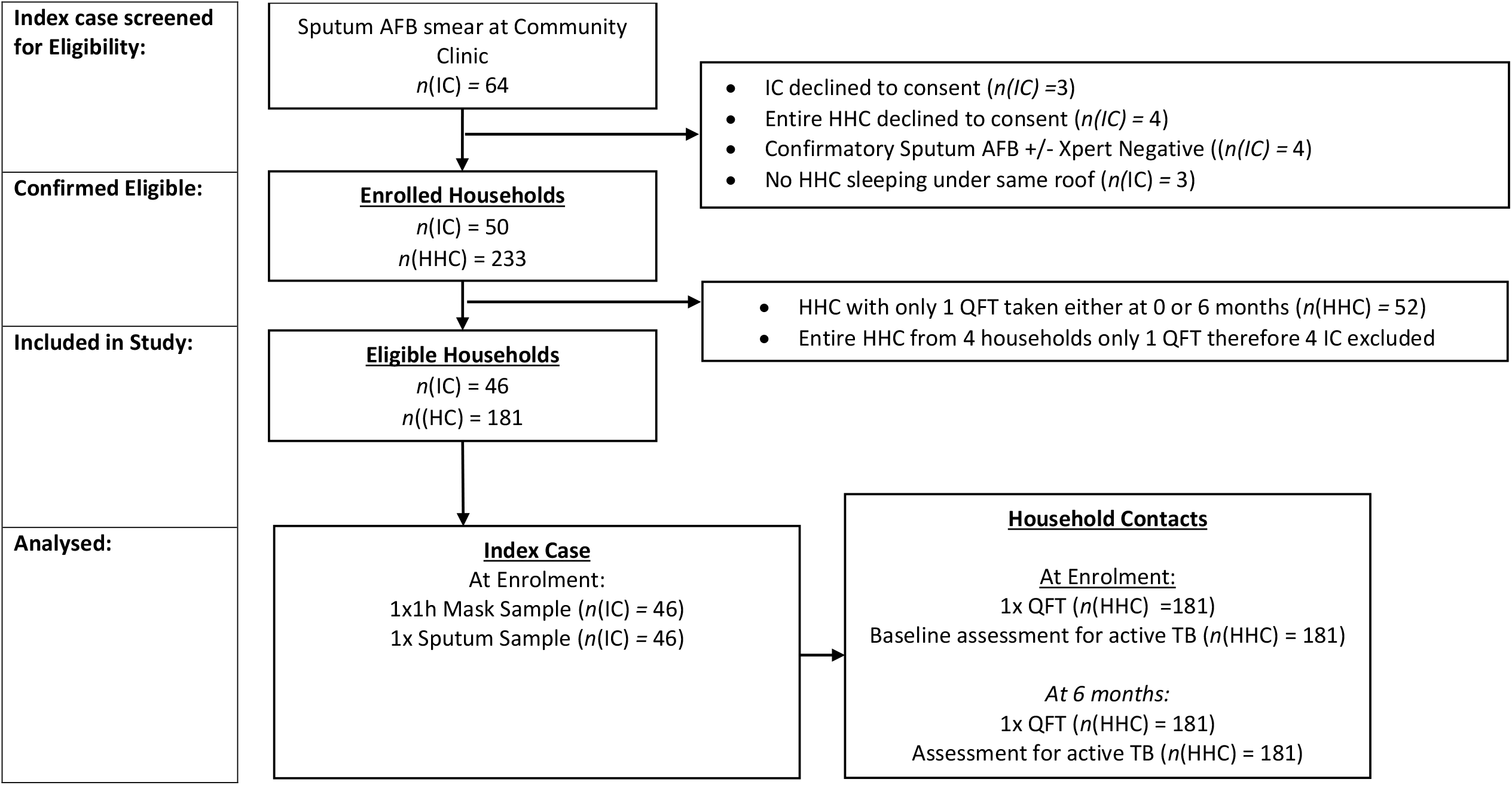
Study profile. AFB – acid-fast bacilli; IC – Index Case, HHC – Household contact, QFT - QuantiFERON TB Gold assay.

### TB cases and Mask Output

All 46 index pTB cases had microbiologically confirmed disease, with 14 (30%) participants having *M. africanum* infection (Table 1). Median age of the cohort was 26 years and 27 participants (59%) were male. HIV co-infection was identified in 2 participants (4%). All participants reported cough and other TB associated clinical symptoms for greater than 3 weeks prior to enrolment and all had moderate or advanced disease on CXR.

**Table 1.** TB case demographic, clinical, radiological and microbiological characteristics, stratified by mask captured IS6110. Mask negative/low positive – IS6110 copies <20,000, Mask high positive – IS6110 copies ≥ 20,000. Values are median (IQR) or n (%) unless otherwise stated ^#^N=18 for cavities present on CXR ^Missing data for MGIT 960 culture: Mask negative/low positive (3); Mask high positive (3); Whole cohort (6)

| Characteristic | Mask Negative/Low Positive | Mask High Positive | Total Cohort | p |
| --- | --- | --- | --- | --- |
| N | 27 | 19 | 46 | 0.43 |
| Age, years | 23 (20-33) | 30 (21-48) | 26 (20-40) | 0.19 |
| Sex |  |  |  |  |
| Male | 17 (63) | 10 (53) | 27 (59) | 0.48 |
| BMI | 18 (17-19) | 18 (17-21) | 18 (17-19) | 0.72 |
| Syx >3 weeks prior to enrolment | 27 (100) | 19 (100) | 46 (100) | 1.0 |
| HIV Status |  |  |  |  |
| Positive | 0 (0) | 2 (10) | 2 (4) | 0.08 |
| CXR findings |  |  |  |  |
| Extent of disease |  |  |  |  |
| Normal | 0 | 0 | 0 | 0.31 |
| Minimal | 0 | 0 | 0 |  |
| Moderate | 4 (15) | 0 | 4 (9) |  |
| Advanced | 23 (85) | 19 (100) | 42 (91) |  |
| Presence of Cavities |  |  |  |  |
| Yes | 8 (30) | 8 (42) | 16 (35) | 0.38 |
| Size of Cavity if present, (cm)<sup>#</sup> | 4 (4-5) | 4 (3-5) | 4 (2-6) | 0.60 |
| Sputum Characteristics |  |  |  |  |
| Acid Fast Bacilli Smear |  |  |  |  |
| Negative | 0 | 0 | 0 | 0.60 |
| 1+ | 8 (30) | 7 (37) | 15 (33) |  |
| 2+ | 10 (37) | 5 (26) | 15 (33) |  |
| 3+ | 9 (33) | 7 (37) | 16 (34) |  |
| Xpert MTB/RIF |  |  |  |  |
| Negative | 0 | 0 | 0 | 0.15 |
| Low | 6 (22) | 3 (16) | 9 (20) |  |
| Medium | 12 (45) | 9 (47) | 21 (46) |  |
| High | 9 (33) | 7 (37) | 16 (35) |  |
| Rifampicin resistance present | 1 (4) | 0 | 1 (2) | 0.40 |
| MGIT 960 culture^ (DTP) | 11 (8-14) | 7 (6-11) | 12 (4-40) | 0.85 |
| Mycobacterial Species |  |  |  |  |
| M. tuberculosis | 21 (78) | 12 (63) | 33 (72) | 0.89 |
| M. africanum | 6 (22) | 7 (37) | 13 (28) |  |

Exhaled Mtb was detected by FMS in 42 (91%) index cases with IS6110 copy numbers varying from 5.3 ×10^2^ to 1.2 ×10^7^ (median 1.8 ×10^4^) among mask positive individuals. A high mask output (>20,000 copies) was detected in 19 (45%) participants.

The proportion with a high mask output did not differ according to measures of sputum bacterial burden, defined by smear AFB (p=0.60) or Xpert MTB/RIF (p=0.15) grades, or time to positive culture with MGIT 960 (p=0.85) (Table 1). There was also no significant differences in CXR severity (p=0.31) or proportion with cavitation (p=0.38) between high and low/negative Mtb mask output groups (Table 1). Index cases with *M. africanum* infection were similarly distributed between the high and low/negative mask output groups (37% vs 22%, p=0.89).

### Household contacts and Prevalent TB infection

The number of household contacts exposed to TB patients with negative, low positive and high positive mask samples were 10, 98 and 73 respectively. No household contacts were found to have active TB at baseline. As the number of household contacts exposed to mask negative patients was small, we combined this group with contacts of index cases having low positive mask samples, for comparative analyses with contacts of high mask output index cases. The groups were statistically matched according to sleeping proximity to their index case (p=0.15), proportions with evidence of previous BCG vaccination (p=0.114) and baseline prevalence of QFT-defined latent Mtb infection (LTBI) (53% for index mask negative/low positive vs 52% for index high mask output contacts, p=0.933) (Table 2).

**Table 2.** Household contacts’ characteristics, stratified by IC exhaled Mtb measured by mask sampling using IS6110. Mask negative/low positive – IS6110 copies <20,000, Mask high positive – IS6110 copies ≥ 20,000. QFT - QuantiFERON TB Gold assay

| Characteristic | Mask Negative/Low Positive | Mask High Positive | Total Cohort | p |
| --- | --- | --- | --- | --- |
| N | 108 | 73 | 181 | 0.46 |
| Age, years | 25 (20-36) | 26 (20-40) | 25 (20-40) | 0.476 |
| Sex |  |  |  |  |
| Male | 39 (36) | 28 (38) | 70 (39) | 0.968 |
| HIV status |  |  |  |  |
| Positive | 1 (1) | 3 (4) | 4 (2) | 0.692 |
| BMI | 22 (19-27) | 20 (19-25) | 21 (19-25) | 0.300 |
| BCG scar |  |  |  |  |
| Present | 57 (53) | 3 (45) | 90 (50) | 0.114 |
| Sleeping proximity to Index case |  |  |  |  |
| Same room | 18 (16) | 14 (19) | 32 (18) | 0.152 |
| Different room, same house | 59 (55) | 50 (68) | 109 (60) |  |
| Same household, different hut/house | 31 (29) | 9 (12) | 40 (22) |  |
| QFT Positivity (%) |  |  |  |  |
| Baseline | 56 (53) | 38 (52) | 94 (52) | 0.933 |
| 6 months | 56 (53) | 44 (60) | 100 (55) | 0.295 |
| IGRA Conversion (Neg-Pos) (%) | 15 (29) | 19 (54) | 34 (39) | 0.043 |
| Quantitate QFT result |  |  |  |  |
| ≥ 1 IU/ml positive | 16 (15) | 25 (34) | 41 (23) | 0.005 |

### Incident Mtb infection

#### Primary outcome measure (QFT conversion)

After excluding household contacts that were QFT positive at baseline, there were 41 (47%) contacts of low or negative mask output and 46 (53%) contacts of high mask output index cases available for assessment of QFT conversion at 6 months. Nineteen QFT conversions (26%) occurred in contacts of high mask output index cases, compared with 15 (14%) in household contacts of low or negative mask output index cases, corresponding with a significantly greater risk of incident Mtb infection in contacts exposed to high mask output index cases (AOR 3.20, 95%CI 1.26 - 8.12, p=0.01, Figure 2, Table S5). The calculated NPV for FMS for incident infection was 75.5% (95% CI: 71.8-85.5).

**Figure 2.**
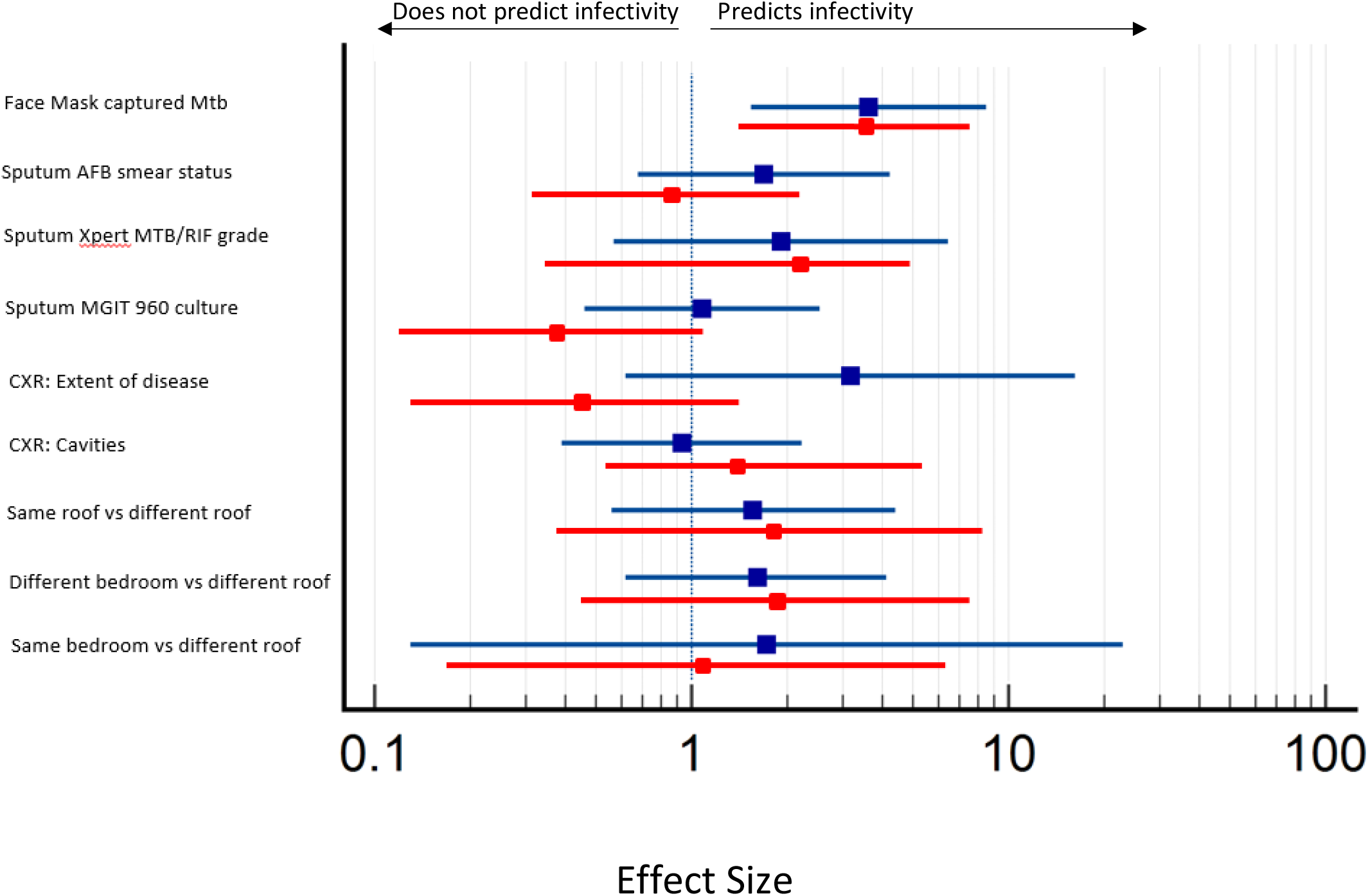
Adjusted Odds Ratios for Predictors of Transmission associated with 2 infection outcome measures; QFT conversion (Red) or QFT change ≥ +1IU/ml (Blue) in exposed Household Contacts.

Measures of sputum Mtb burden (AFB grade, Xpert grade) radiological extent of disease and sleeping proximity showed no significant association with QFT conversion in our models (Table 3). However, a possible association between DTP in MGIT culture and QFT conversion that approached statistical significance was observed (p=0.07) (Figure 2, Table S5).

#### Secondary Outcome measure: IFN-ƴ increase of ≥ 1 IU/ml at 6 months

Applying this criterion to the full cohort of 181 household contacts, 25 (34%) new or recently acquired Mtb infections occurred in contacts of high IS6110 mask output TB cases, compared with 16 (15%) Mtb infections in contacts of low positive/negative IS6110 index cases. Contacts of TB cases with a high IS6110 mask output had a greater than threefold increased risk of incident Mtb infection than contacts of a negative or low positive IS6110 mask (0R 3.62, 95%CI 1.54 - 8.53, p=0.003, Table 4). The calculated NPV for FMS for incident infection was 80.5% (95% CI: 73.4-86.0).

As for the primary outcome measure, sputum Mtb burden, radiological disease and sleeping proximity were not statistically associated with incident Mtb infection in household contacts (Figure 2, Table S5). Finally, we observed no difference in the proportion of contacts with newly acquired infection defined by either criterion, according to the bacterial strain of the index case (QFT conversion: p=0.28; IFN-ƴ ≥1IU/ml: p=0.22).

## Discussion

Our data demonstrate that FMS provides a simple and non-invasive tool to support the clinical assessment of pTB in a high burden setting, for diagnosis and stratifying risk of TB transmission to household contacts more effectively than indices of infectivity widely applied in clinical practice.

Specifically, FMS detected exhaled Mtb in a large proportion (91%) of the index pTB cohort and household contacts of high Mtb output index cases had a greater than threefold increased likelihood of incident Mtb infection based on QFT conversion after 6 months. This stratification of risk was not demonstrated with either sputum bacillary burden or radiological extent of disease, which are indices currently used by clinicians for this purpose^23^.

Consistent with previous reports, we also found no difference in disease severity, FMS mycobacterial output or transmission in contacts associated with pTB caused by *M. africanum*, in the 28% of our index cohort infected with this lineage^24^. Our findings are strengthened by conducting the study, using the robust TB Case-Control (TBCC) platform based at the Medical Research Council Unit, which enabled systematic longitudinal evaluation of the recruited cohorts^25^.

Transmission studies are limited by the absence of a reliable objective biomarker for this outcome. We used QFT seroconversion as our primary outcome measure of transmission. This is the most widely used method for identifying incident infection with Mtb complex and presumes transmission from the index case following recent close contact. Validity of this approach to detect recently acquired Mtb infection is supported by studies that have demonstrated increased risk of progression to TB, an outcome associated with recent infection, compared with subgroups that have a persistently positive serial QFT result^20,26,27^. However, attribution of incident infection to transmission is inferred; furthermore although specific, QFT conversion is likely to capture only a subset of transmission events as exposed contacts may have acquired infection prior to diagnosis of the index, or from another source in high TB burden settings.

In this study, only 87 (48%) of household contacts could be included to primary outcome analysis; this compares with 27% of the household contacts in the CASS transmission study^8^ and emphases the need for a more accurate measures of transmission. To address this, we included a significant quantitative increase in QFT^18–21^ as a secondary outcome measure of transmission, allowing inclusion of our complete household contact cohort in analyses. The consistency of results between these two measures of transmission support consideration of quantitative changes in IGRA response for future studies.

As a tool for measuring exhaled bacilli, FMS is comparable with aerosol sampling methods. In this respect, there are notable similarities and differences in outcome between our study and reported studies using CASS. Both FMS and CASS output associate significantly with incident Mtb infection in household contacts, but this is not observed for traditional markers of infectivity, including sputum bacillary burden and radiological extent of disease^7–9,28–30^. Furthermore, our previous studies with FMS identified inconsistencies in the relationship between mask and sputum bacillary burden that have also been reported by aerosol studies^8,12,29,31–33^. Together these observations support the view that exhaled and aerosolised Mtb constitute a distinct Mtb pool that associates with transmission more strongly than other measures of bacillary burden^30^.

In this respect, we note some important differences between FMS and CASS. Firstly, there is a significant difference between the methods in the proportion of Mtb positive individuals identified that may influence the stratification of transmission risk. In this study Mtb was detected in 91% of pTB patients sampled with FMS, compared with 45% of patients in a similarly designed transmission study using CASS^8^. In that study the authors reported a greater than 9 fold increased odds of IGRA seroconversion in household contacts of CASS positive patients with a high aerosol output, compared with the three-fold difference in odds that we have identified using FMS. However, 36% of the contacts of index cases with a negative CASS and 47% of the contacts of cases with a low Mtb CASS output had evidence of recent transmission. In contrast, the present study found only 14% of contacts of negative or low FMS Mtb output pTB cases had QFT conversion. Together these data suggest that while aerosol Mtb output measured with CASS is more specific, it is considerably less sensitive than FMS for informing transmissibility of infectious pTB. From a clinical translational perspective. Furthermore, the calculated NPV of FMS at 79.5% and 80.5% indicates the possibility that FMS could be used to focus contact follow up in resource limited settings.

These differences between FMS and CASS may reflect the difference in sampling type. CASS only captures culturable bacilli and is therefore likely to collect a much smaller proportion of Mtb expelled by individuals than mask sampling. By capturing DNA, mask sampling captures a wider range of bacillary states, including free DNA and non-replicating cells, with a potentially lower but clinically relevant transmission potential.

This study has some limitations. We were not powered to reliably assess the association between the existing clinical measures of infectivity and transmission. Although no association was demonstrated in this study, it is possible that significant associations would be identified in a larger cohort study. Our finding of a near significant association between DTP in MGIT culture and QFT conversion would support this view, and a previous larger cohort study did identify an association between contact sleeping proximity and Mtb transmission^34^ that was not observed here. Nevertheless, the consistency of our findings with previous aerosol transmission studies would indicate the greater importance to transmission of the Mtb population captured in exhaled air.

Our method for measuring transmission is a flawed gold standard and improved measures of transmission need to be developed to support future transmission studies. We also acknowledge that the lack of a population control and the 6 month follow-up period used in this study, which is longer than other transmission studies^8,11,35^, does introduce further uncertainty about the origins of any QFT response, particularly in a high TB burden setting. Furthermore, we did not perform FMS in the contact cohort and although our assessments did not identify any cases of clinically active TB, we are unable to determine whether any contacts developed transmissible subclinical TB.

Finally, the relatively low HIV-TB co-infection rate in The Gambia makes these results difficult to generalise for many other high burden TB areas, since the influence of HIV on both index case infectivity and susceptibility of contacts can potentially alter the findings. However, as a tool to support pTB diagnosis, the performance of FMS in this study is comparable with the proportions of patients having a positive Mtb output in our previous published work that was predominately in an HIV positive cohort. ^12^. Further work in high HIV high TB burden and low TB burden settings is needed to fully assess the generalisability of mask sampling as a clinical infectivity tool.

In summary, face mask sampling is a non-invasive, inexpensive and easily deployable tool that demonstrates capability to stratify transmission risk from individuals with pulmonary TB. This builds on our previous work supporting a role for mask sampling as a highly sensitive diagnostic tool^12^. Together, these studies support the potential of FMS as a clinical tool to enhance TB control programmes necessary for eradication of TB^36^ and provides an epidemiological tool to better characterise Mtb transmission within complex community settings.

## Data Availability

All data produced in the present work are contained in the manuscript

