## Supplemental data for "Exhaled Mycobacterium tuberculosis predicts incident infection in household contacts"

Supplementary data

### Study Methods

#### Study Setting

This study was conducted in the Greater Banjul Area (GBA) of The Gambia using the TB Case-Control (TBCC) platform based at the Medical Research Council Unit The Gambia at LSHTM[1]. The GBA is a mixture of urban, peri-urban and rural dwellings where the majority of all the TB cases in The Gambia are notified[2]. Many of the households in The Gambia still live in compounds, which comprise of multiple small sleeping huts arranged around a central communal area used for cooking, eating and socialising.

#### Screening and recruitment

To identify index cases (IC), the results of TB diagnostic sputum samples processed at community clinics were screened daily by the study team. Patients with positive sputum samples who fulfilled the eligibility criteria were approached to participate in the study by a fieldworker prior to their return to clinic for treatment. They were given written information about the study which was translated into their preferred language. Patients who were willing to participate were asked to sign a consent form which was countersigned by an interpreter or witness if one was involved in the consent procedure.

Following standard practice, the fieldworker also introduced the TBCC to their household contacts (HHC) and enquired about their interest in being recruited. Both a screening and enrolment log was kept of all patients assessed and approached.

Inclusion Criteria

The following inclusion criteria were used to assess the index cases:

- Aged ≥ 18 years.
- AFB+ sputum sample taken at the community clinic.
- Yet to commence TB treatment.
- HHCs willing to consent into TBCC platform.
- At least one HHC sleeping under the same roof
- Able to give genuine informed consent.

Exclusion Criteria

ICs meeting the following criteria at baseline were excluded from the study:

- Requiring oxygen therapy.
- Unable to understand or comply with sampling instructions given to them by the study team due to medical condition.
- No HHCs enrolled into TBCC.

#### Sampling methods

All sampling from ICs was undertaken at a single clinic visit prior to starting TB treatment. Collected samples were stored and transported at +4°C and sample collection, transportation and processing logs were kept ensuring they were processed within 72 hours of collection.

**Index TB participant Mask and Sputum Sampling**

Each index TB participant wore a modified FPP1 face mask containing a gelatine filter (pore size 0.3µm, Sartorius, Germany. Figure S1). Subjects were specifically not required to perform any vocal manoeuvres and could to cough, talk, laugh or sleep as desired. If they needed to expectorate then they were asked to lift the mask briefly, after coughing, to expectorate into a sputum collection pot.

Each participant underwent mask sampling for an hour. Subjects were observed to ensure that the mask was worn for the whole hour of sampling and participant behaviour was recorded, including anytime the mask was directly in front of the mouth. Sleep was documented if the participant had their eyes closed for >10 minutes and were not obviously rousable by noise, resting was noted if the participant had their eyes closed for <10 minutes at a time and/ or had their eyes open but not engaging in any activity such as talking, eating, reading etc. Other activities such as eating, washing, reading and talking were documented.

If an index TB participant did not spontaneously expectorate sputum during mask sampling, they were asked to produce one at the end of sampling.

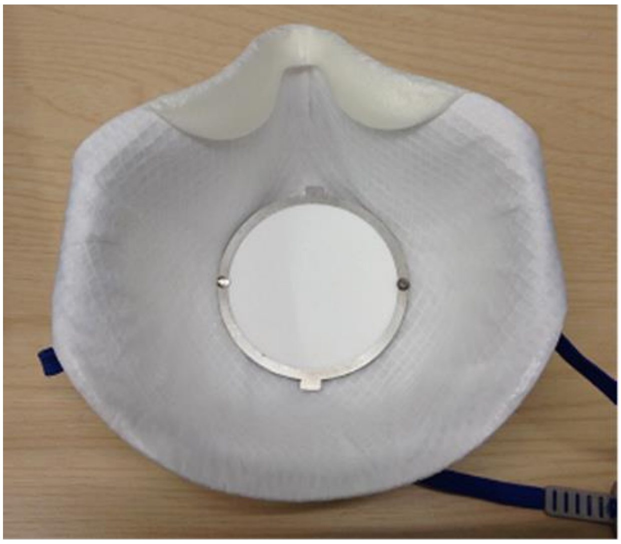

**Figure S1: FPP1 face mask containing a gelatine filter with a diameter of 60mm**

**Data Collection**

The TBCC platform routinely collects clinical and demographic data for each person enrolled as an IC or HHC at baseline and six months. Paper copies of laboratory CRFs were designed in Microsoft Word and Excel. The TBCC team was trained on data collection and entry by the author prior to the start of the study to ensure standardised documentation of observations and sample collection. All paper CRFs were stored in the MRC Unit's research archive and will be retained for 5 years after study completion. Clinical and demographic data were stored on the electronic TBCC database and a smaller database was created for analysis of participants enrolled in this study.

Data collection for the Index Case (IC)

At enrolment, ICs underwent a detailed medical history and clinical examination to assess disease severity. CXRs were taken and sputum analysis, including sputum AFB smear microscopy (at the MRC Unit), Xpert MTB/RIF, spoligotyping and liquid culture were performed (BACTEC MGIT 960).

Radiological changes were visually assessed on a standard full sized postero-anterior CXR using an in-house method which all members of the research team involved in CXR interpretation had been trained to use. CXR grading criteria are defined in table S1. CXR for the ICs were validated independently by the author who was blinded to the initial TBCC assessment of the CXR.

| Grade | Criteria observed on CXR |
|---|---|
| Normal | No abnormality seen |
| Minimal Disease | Infiltrates in 1-2 lung zones without cavitation |
| Moderate Disease | Infiltrates in 3+ lung zones without cavitation |
| Severe Disease | Presence of cavitation and/or all lung zones |

Table S1. In- house grading of abnormalities seen on CXR of participants in study.

ICs were then followed up regularly during their treatment for TB and then every 6 months for 2 years.

**Data collection for the Household contact**

Each HHC enrolled into the TBCC was assessed at baseline for signs of active TB. This included a detailed medical history and clinical examination, with CXR and where possible sputum analysis. Those with active disease were offered treatment and followed regularly throughout treatment, whereas those without active disease underwent HIV and IGRA testing.

HHC were followed up again at 6 months, assessed for active TB disease and if negative underwent IGRA testing. If HHC developed TB symptoms during this 6 month period they were reviewed by the TBCC team and again assessed for active disease.

Data was collected on the whole household, including the number and age of all people living in each compound (regardless of number enrolled in TBCC) and sleeping proximity of each person to index case. HHCs were then stratified into the following groups, according to sleeping proximity to the IC: sleeping in same room as IC, sleeping under the same roof as the IC but not in the same room, and sleeping under a different roof.

**Sample Processing**

**Mask samples**

Mask samples were processed using the methods detailed previously[3]. Briefly, gelatine filters were dissolved in 1.5mls of 2% w/v NaOH and incubated at room temperature for 15 minutes before neutralising with 190μl 4M

HCL. Samples were agitated by hand at 0 and 8 minutes. The dissolved filter was then centrifuged at 13,400 x*g* for 10 minutes, the supernatant removed and the pellet overlaid with 100µl of TE buffer (20mM Tris and 2mM EDTA pH 8.0), prior to storage at -80°C. Bacterial pellets an in-house extraction method modified from that outlined by Reddy and colleagues[4]. 100µl Chelex-NP40 (50% w/v Chelex-100, 1% w/v Non-idet P40, 1% w/v Tween 20) was added to the defrosted bacterial pellet along with 0.3g glass beads (150-212µm Sigma-Aldrich USA). The sample was then homogenised in a Fast Prep at 6.5 m/s for 45 seconds and ice incubated for 5 minutes. This homogenisation step was repeated 3 further times prior centrifugation at 13,400xg for 2 minutes. 200µl of the supernatant was heated at 95°C for 30 minutes prior to removal from the CL3.

Copy numbers of IS6110 were quantified in each sample by real-time q-PCR run on a Rotor-Gene (Qiagen UK) using a TaqMan IS6110 assay outlined by Akkerman and colleagues as 'in-house TaqMan -10' [5]. Real-time PCR signals were analysed by Rotor-Gene 6000 Series Software 1. The functions "slope correct" and "ignore cycles" were applied to analyses. In accordance with Dorak and colleagues only runs with correlation coefficients ($R^2$ ) and reaction efficiencies above 0.99 and 0.8 respectively were included for analysis[6]. Technical replicates were undertaken in triplicate and considered reproducible if the difference in cycle threshold (Ct) was less than 1.

**Sputum processing**

The sputum sample collected underwent the following routine microbiological investigations as part of the TBCC platform:

**Xpert MTB/RIF**

1ml of sampled sputum was removed prior to decontamination for Xpert MTB/RIF processing. Following the manufacturer's instructions, the sputum aliquot was mixed with the Xpert buffer and incubated at room temperature prior to loading into the Xpert MTB/RIF machine.[7]

**Microscopy and Decontamination of Sputum**

Sputum samples were all initially screened for the presence of AFB by Auramine microscopy. Fresh samples were decontaminated by the NALC-NaOH method as described by Peres and colleagues.[8] The purity of all decontaminated samples was subsequently checked on blood agar for 48h at 37°C and screened for AFB by Ziehl-Neelsen staining during which time the decontaminated sputa were stored at -20°C, prior to inoculation.

Bacterial Culture

Confirmed blood agar negative and AFB positive samples were cultured within the BACTEC MGIT 960 System (MGIT 960; Becton Dickinson Microbiology Systems, Sparks, Maryland, USA) at 37°C according to the manufacturer's instructions. Instrument positive vials were removed from the machine and again subjected to purity check using blood agar and ZN microscopy to confirm the presence of AFB.

**Spoligotyping**

Aliquots of the MGIT cultures were taken and heat killed. These lysates were genotyped by spoligotyping as described by Kamerbeck *et al.*[9] Participants' isolates were assigned to specific TB lineages using the TB-lineage tool within the TB-Insight public database.[10]

**QuantiFERON analysis**

The tubes were transported at ambient conditions to the immunology laboratory at MRC Fajara and incubated overnight (18 to 22 hours) at 37 °C, 5% $CO_2$. Tubes were then centrifuged at 2,500 x g for 15min and supernatants harvested, aliquoted and frozen (−80 °C) until use. QFT ELISA was performed in bulk according to manufacturer's instructions and Interferon- γ (IFN-γ) levels determined using QFT GIT software version 2.6.2.

**Statistical Analysis**

Data were analysed using GraphPad Prism (Version 7, USA) Excel 2010 (Microsoft, USA) SPSS Version 22 (IBM, USA) and STATA Version 13.1 (StataCorp LP, USA).

Baseline characteristics were described for all ICs and HHCs. Mask Mtb output (measured using IS6110) were found to be non-normally distributed using the Shapiro-Wilk test. Univariate analysis of demographic and clinical features of the ICs were undertaken using non parametric tests; for correlations between continuous variables Spearman's rank test was used, for comparison between continuous and categorical variables the following tests were used: Mann Whitney U test if the categorical variable had 2 groups, Kruskal-Wallis test if > 2 groups. For comparison of categorical data groups, a Chi square test was used if 2 groups or Cramer V if > 2 groups. A p-value of <0.05 was considered statistically significant.

**Comparison of excluded and included participants**

| Characteristic | Excluded | Included | *p* |
|---|---|---|---|
| **N** | 4 | 46 | |
| **Age, years** | 28 (27-59) | 26 (20-40) | 0.33 |
| **Sex** | | | |
| Male | 3 (75) | 27 (59) | 0.16 |
| **BMI** | 18 (15-18) | 18 (17-19) | 0.47 |
| **Syx >3 weeks prior to enrolment** | 4 (100) | 46 (100) | 0.78 |
| **HIV Status** | | | |
| Positive | 0 (100) | 2 (4) | 0.90 |
| **CXR findings** | | | |
| **Extent of disease** | | | |
| Normal | 0 | 0 | 0.78 |
| Minimal | 0 | 0 | |
| Moderate | 0 | 4 (9) | |
| Advanced | 4 (100) | 42 (91) | |
| **Presence of Cavities** | | | |
| Yes | 1 (25) | 16 (35) | 0.87 |
| Size of Cavity if present, (cm)* | 3 (3) | 4 (2-6) | 0.23 |
| **Sputum Characteristics** | | | |
| **Acid Fast Bacilli Smear** | | | |
| Negative | 0 | 0 | 0.62 |
| 1+ | 1 (25) | 15 (33) | |
| 2+ | 1 (25) | 15 (33) | |
| 3+ | 2 (50) | 16 (34) | |
| **Xpert MTB/RIF** | | | |
| Negative | 0 | 0 | 0.39 |
| Low | 1 (25) | 9 (20) | |
| Medium | 0 | 21 (46) | |
| High | 3 (75) | 16 (35) | |
| Rifampicin resistance present | 0 | 1 (2) | 0.96 |
| **MGIT 960 culture^ (DTP)** | 10 (6-16) | 12 (4-40) | 0.72 |
| **Mycobacterial Species** | | | |
| *M. tuberculosis* | 3 (75) | 33 (72) | 0.87 |
| *M. africanum* | 1(25) | 13 (28) | |

**Table S2. TB case demographic, clinical, radiological and microbiological characteristics, for those included and excluded from the study.** Values are median (IQR) or n (%) unless otherwise stated *N=18 included cohort, N=1 excluded cohort ^Missing data for MGIT 960 culture: Included cohort (6), excluded cohort (0)

| **Characteristic** | **Excluded** | **included** | ***p*** |
|---|---|---|---|
| **N** | 52 | 181 | |
| **Age, years** | 26 (17-64) | 25 (20-40) | 0.78 |
| **Sex** | | | |
| Male | 27 (52) | 70 (39) | 0.09 |
| **HIV status** | | | |
| Positive | 1 (2) | 4 (2) | 0.90 |
| **BMI** | 22 (20-25) | 21 (19-25) | 0.12 |
| **BCG scar** | | | |
| Present | 29 (56) | 90 (50) | 0.06 |
| **Sleeping proximity to Index case** | | | |
| Same room | 12 (23) | 32 (18) | 0.20 |
| Different room, same house | 24 (46) | 109 (60) | |
| Same household, different hut/house | 16 (30) | 40 (22) | |
| **QFT Positivity (%)*** | | | |
| Baseline | 23 (44) | 94 (52) | 0.63 |
| 6 months | 1 (50) | 100 (55) | 0.90 |

**Table S3. Household contacts' characteristics, for those included and excluded from the study.** QFT - QuantiFERON TB Gold assay * Missing data for QFT positivity: Baseline excluded (11) included (0); 6months: excluded (50) included (0).

**Defining outcome measures**

The main outcome for this study was Mtb infection in household contacts. This was investigated in several ways. The primary outcome measure was IGRA conversion in household contacts with those HHC who were IGRA positive at baseline excluded from the analysis. Here conversion was defined as an Interferon-$\gamma$ level of <0.35 IU/ml at baseline and then ≥0.35IU/ml at 6 months. Removing HHCs with a positive IGRA at baseline (and therefore only counting IGRA conversions observed since diagnosis of the IC) reduces the risk of confounding due to infections in HHCs that are unlinked to the IC. However in TB endemic areas, such as The Gambia the prevalence of an IGRA positive result is ~50% so many HHCs will be excluded from analysis.

The IGRA test is not just binary and there has been recent interest in trying to understand the quantitative result of an IGRA test and seeing if this related to infection. By using the quantitative change between the IGRA reading at baseline and at 6 months (the delta change) the whole cohort could be included which would increase the confidence in the results seen. Studies have confirmed that change in the quantitative value of IGRA is associated with a higher incidence of TB disease progression, [11] having a higher Interferon-ɣ result increased the likelihood of progression to TB disease in contacts of pulmonary TB [12] and levels of Interferon-ɣ are dynamic and reversion is common in TB endemic areas. [13]

Studies have also highlighted that an Interferon-ɣ level of < 1 IU/ml is less likely to represent a significant infection event [14,15]. Therefore we investigated a second outcome measure, an increase in Interferon-ɣ of ≥ 1 IU/ml regardless of baseline Interferon-ɣ result.

**Model building for the association of Index and HHC characteristics with Transmission**

To allow for clustering effects relating to clinical and epidemiological characteristics of the index case and HHC, factors associated with TB infection were determined using an unadjusted multilevel mixed-effects logistic regression model at a 10% significance level. The multivariable models were built using a forward selection of significant univariate factors, only including factors significant at the 10% level in the multivariable model (Table S2).

**Confounders in outcome model**

Using IGRA conversion in the HHC as the outcome measure, age of the HHC was significantly associated (p=0.04); and using an Interferon-ɣ quantitative increase of ≥ 1IU/ml in the HHC as the outcome measure, the presence of a BCG scar in the HHC was significant (p=0.04).

| Predictor | Outcome measure (OR [95% CI](p Value)) | |
|---|---|---|
| | IGRA Conversion | ≥ 1 IU/ml increase in IFN-ɣ |
| **Index** | | |
| CXR disease | 0.49 [0.12, 1.93] (0.30) | 3.45 [0.70, 17.14] (0.12) |
| Cavities present | 1.73 [0.58, 5.13] (0.31) | 0.97 [0.42, 2.25] (0.94) |
| HIV status | Numbers too small to undertake analysis | |
| BMI | 1.07 [0.90, 1.27] (0.46) | 1.04 [0.91, 1.18] (0.57) |
| Strain | 0.69 [0.24, 1.97] (0.47) | 0.60 [0.26, 1.41] (0.23) |
| **Household Contact** | | |
| Age | 1.04 [1.00, 1.07] (0.04) | 0.99 [0.96, 1.02] (0.58) |
| Sex | 0.83 [0.31, 2.21] (0.70) | 1.26 [0.58, 2.71] (0.55) |
| Proximity | 2.02 [0.39, 10.30] (0.39) | 1.24 [0.46, 3.30] (0.67) |
| | 1.76 [0.29, 10.85] (0.53) | 1.36 [0.41, 4.49] (0.61) |
| BCG | 1.79 [0.38, 8.40] (0.45) | 2.34 [1.02, 5.36] (0.04) |
| HIV status | Numbers too small to undertake analysis | |
| BMI | 1.02 [0.92, 1.13] (0.75) | 0.98 [0.91, 1.06] (0.58) |

Table S4 Univariable analysis of Household contact and Index characteristics as predictors for TB infections

**Effect size analysis of IS6110**

The primary exposure variable was the quantification of Mtb DNA captured from the IC using mask sampling.

To evaluate a relationship between number of copies of IS6110 and IGRA conversion in household contacts (primary outcome measure), mask output was divided into 2 groups after first looking at the trend in risk of IGRA conversion by finely divided categories of genomic copies. Final groupings for IS6110 were (1) ≥20,000 copies or (2) <20,000 copies or negative. These cut-offs were chosen because this was the point at which an increase in IGRA conversion risk was noted (see Figure S2). The methods for this effect size analysis are based on the published methods by Jones-Lopez *et al* using CASS.[16]

| Household contact infection predictor | QFT conversion (negative to positive) | | | QFT change ≥ +1 IU/ml | | |
|---|---|---|---|---|---|---|
| | AOR | 95% confidence interval | p value | AOR | 95% confidence interval | p value |
| **Face Mask captured Mtb[¥]** | 3.20 | 1.26 - 8.12 | **0.01** | 3.26 | 1.54 - 8.53 | **0.003** |
| **Sputum Mtb Burden** | | | | | | |
| AFB smear status* | 0.82 | 0.30 – 2.20 | 0.69 | 1.69 | 0.68 - 4.22 | 0.26 |
| Xpert MTB/RIF grade[#] | 1.11 | 0.33 – 3.73 | 0.86 | 1.92 | 0.57 – 6.45 | 0.29 |
| MGIT 960 culture^ | 0.38 | 0.13 – 1.09 | 0.07 | 1.08 | 0.46 – 2.54 | 0.85 |
| **Chest Radiograph changes** | | | | | | |
| Extent of Disease | 0.49 | 0.13 - 1.79 | 0.28 | 3.17 | 0.62 – 16.22 | 0.17 |
| Presence of Cavities | 1.45 | 0.52 - 4.01 | 0.48 | 0.93 | 0.39 – 2.23 | 0.88 |
| **Sleeping Proximity of HHC** | | | | | | |
| Same roof (bedroom or house) vs different roof | 1.75 | 0.38 – 8.03 | 0.47 | 1.56 | 0.56 – 4.39 | 0.39 |
| Different bedroom same roof vs different roof | 1.84 | 0.43 – 7.81 | 0.41 | 1.61 | 0.62 – 4.13 | 0.32 |
| Same bedroom vs different roof | 1.06 | 0.17 – 6.82 | 0.95 | 1.72 | 0.13 – 22.91 | 0.68 |

**Table S5. Adjusted Odds Ratios for Predictors of Transmission associated with 2 infection outcome measures; QFT conversion or QFT change ≥ +1 in exposed Household Contacts.** [¥]IS6110 - ≥20,000 copies vs <20,000 copies or negative, *AFB – 1+ vs >1+, [#]low vs >low, ^ Days to Positivity 0-10 vs >10 days and negative
